# Validation of the Brazilian version of the Processes of Change Questionnaire in Weight Management in adults with overweight and obesity in Brazil

**DOI:** 10.1101/2023.03.20.23287450

**Authors:** Quênia de Carvalho, Paola Rampelotto Ziani, Bruno Braga Montezano, Jeferson Ferraz Goularte, Adriane R Rosa

## Abstract

**Introduction:** This study aimed to analyze the psychometric properties of the Brazilian version of the Processes of Change Questionnaire in Weight Management (P-Weight) questionnaire.

**Methods:** A total of 656 adults participated in the study, including people in weight loss treatment and people from the general community. All participants responded to the Process of Change Questionnaire (P-Weight), the Stages of Change Questionnaire (S-Weight), and the Eating Attitudes Test (EAT-26), which assesses risk of eating disorder used as a measure of external validity. Socio-demographic variables were also investigated.

**Results:** The 33-item P-weight questionnaire showed satisfactory psychometric properties with high internal consistency (Cronbach’s alpha=0.959). Exploratory and confirmatory factor analyses revealed a 4-factor model similar to the original Spanish version of P-Weight with a slightly rearrangement of the items (KMO=0.92, df (528, n=328) =8,401.015; p<0.0001). We found significant associations between processes and stages of change (p<0.001) and a moderate correlation between the four subscales of P-weight and EAT-26 (p<0.001). Finally, the mean score of P-Weight were higher in clinical sample compared to the general community, suggesting the sensitivity to discriminate cases and controls (p<0.001).

**Conclusion:** This study showed the validity and reliability of the Brazilian version of the P-Weight scale. Therefore, the P-Weight is readily available to help professionals to employ precision interventions to weight loss considering the patient’s motivational stage in combination with their individual use of the cognitive processes of change.

## Introduction

In Brazil, the percentage of adult people who living with overweight is 57.5% for men and 62.6% for women [1], which suggests that obesity is a public health problem. The long-term consequences of obesity have revealed increased risk for other primary lifestyle diseases, including coronary heart disease, hypertension, stroke, type 2 diabetes mellitus, sleep apnea, osteoarthritis, prostate disease, autoimmune conditions, neuropsychiatric disorders, and others [2]. Current approaches emphasize that lifestyle changes with the adoption of low-energy dietary patterns are effective in reducing body weight and may prevent obesity-associated chronic diseases [3]. However, most people living with obesity do not stay on weight loss programs long enough and only a few of them lose and maintain the ‘new’ weight [4]. Thus, a study (n=50,081) showed that 95% of people who try to lose weight, regain it in a period of five years after a dietary, pharmacological, or behavioural intervention; the faster the weight loss, the faster the weight recovery; with this behaviour more evident in cases than control groups [5]. Therefore, the long-term success of weight loss programs depends on the ability of individuals living with obesity to change their behaviours, which is extremely difficult for most people, especially regarding to maintain a new diet pattern and to exercise regularly [6].

Considering behaviour as the key to weight loss, The Transtheoretical Model (TTM) proposed by James Prochaska in 1979, has been used in many fields of Medicine, including obesity [7]. TTM is based on behaviour change considering the individual’s readiness to change and the process of change [8]. According to the TTM, stages of change represent the temporal, motivational, and constancy aspects of change that focus on ‘when’ people change [4], indicating that the individual moves through these stages when modifying their behaviour. The stages of change can be modelled in five stages: 1) Pre-contemplative (PC), individual does not want to make any change; 2) Contemplative (C), individual is ambivalent about change; 3) Preparation (P), individual begins to plan and commit to change; 4) Action (A), individual practices actions aimed at change; and 5) Maintenance (M), individual works to maintain and sustain change for a long time [9]. On the other hand, the processes of change refer to “how” people change and consist of activities in which individuals engage when trying to modify complex behaviours [4]. Although the TTM has been initially applied to smoking cessation attempts [10], it has been applied against obesity with relative success [11]. Furthermore, processes of change have been identified as predictors of behaviour change in interventions aimed at health promotion, such as quitting smoking, healthy eating behaviour, physical exercises, and weight control [4].

The identification of stages and processes of change is critical to individualize interventions and may help to increase the treatment effectiveness. For that, specific questionnaires to assess stages and processes of change in weight management have been proposed by the scientific community [12]. The Stages of Change Questionnaire (S-Weight) and Process of change questionnaire in weight management (P-Weight) seem to be effective to assess the readiness for change and the processes faced during change in patients who are controlling their weight [13]. Both instruments were validated in populations of Spain [4] and United Kingdom (UK) [14] with acceptable psychometric properties, which allow their use to implement behaviour change interventions for weigh loss. When applied in individuals living with obesity on the waiting list for bariatric surgery, P-weight and S-weight showed they were in the Action and Maintenance stages for change. There was also relationship between stages for change and the body mass index (BMI), since the BMI is lower in individuals who are in the Action and Maintenance stages, compared to those who reported being in the Pre-Contemplation stage for change [6]. Therefore, the identification of these stages and processes of change may contribute to weight loss and increased postoperative success rates.

Using the P-Weight and S-Weight scales, A Thai randomized clinical trial that aimed at determining the effects of individualized nutrition counselling, combined with a TTM model on weight loss in people with overweight and obese found that individuals who received the combined nutritional counselling with TTM experienced greater weight loss than the control group, which received only an educational handbook [15]. Another recent study evaluated the stages and processes of change for weight loss in patients living with obesity who had a recent stroke. It showed that patients who are emotionally invested in weight loss and take action regarding their weight are more likely to be in an active phase of behaviour change for weight loss. Although studies using the P-Weight and S-Weight scales have shown promising results, a Cochrane review reported that the effectiveness of TTM in promoting weight loss is still unclear due to the small sample and heterogeneity in the methodologies used, making it difficult to draw effective conclusions [10].

Thus, considering the high prevalence of overweight and obesity in Brazil and the absence of validated scales capable to assess stages and processes of changes in individuals with overweight and obesity, the aim of this case-control study was: 1) to analyze the psychometric properties of the transculturally adapted Brazilian version of the P-Weight; 2) to assess the relationship between stages and process of change; and 3) to assess the relationship between P-Weight and external measures regarding eating disorders such as EAT.

## Materials and Methods

### Translation, transcultural adaptation, and content validity

Initially, the P-Weight scale was independently translated from Spanish to Portuguese by two bilingual translators (QC and ARR) and then the back translation was done by a native speaker of Brazilian Portuguese (DB) that was unaware of the study objectives. Then, the adaptation of semantic and idiomatic equivalences was carried out by the responsible researchers (QC, JG and ARR). In the next step, the instruments were administered to the ten specialists (clinical nutritionists) to detect possible divergences in the understanding of the item by Brazilians. In addition, each item was evaluated regarding cultural equivalence. All considerations made by specialists were reviewed by our research team. Then, a final Brazilian Portuguese version was obtained and applied to a sample of ten patients who were undergoing weight loss treatment to verify its applicability.

The P-Weight is composed of 34 items to measure the processes of behaviour change. For each question, by a 5-point Likert scale, the person scores according to their level of agreement for the question. The P-Weight suggests that four processes of change are involved in this setting: i) emotional re-evaluation (EmR; emotional reactions to being overweight and what will happen if they engage in weight management actions); ii) weight consequences evaluation (WCE; the individual’s awareness of consequences that overweight has on their life, becoming aware that actually they have a weight problem); iii) weight management actions (WMA; those specific actions that people engage when trying to manage their weight); iv) environmental restructuring (EnR; actions aimed to modify the individual’s environment to promote weight management) [16]. The scores for each of the four change processes are calculated by adding the scores obtained on the items belonging to the same subscale. None of the items are reverse scored. To make the scores of the different subscales comparable, these scores are transformed into a scale from 0 to 100, with 0 being the non-use of a given process of change, and 100 being the full use of such process [16]. The highest use of a process is represented by scores above 50, whereas the lowest use of a process, by scores below 50 [15].

### Sample, inclusions criteria, and recruitment

A cross-sectional observational study was carried out using an anonymous online questionnaire. The sample was non-probabilistic by convenience (n=656), composed of people from the general community (community sample) and participants in weight loss programs at the Hospital das Clínicas de Porto Alegre (HCPA) and private clinics in Porto Alegre (clinical sample). The inclusion criteria for the community sample were individuals aged over 18 years and BMI above 18.5 kg/m^2^; and the inclusion criteria for the clinical sample were individuals aged over 18 years, BMI above 18.5 kg/m^2^, and participation in nutritional or pharmacological treatment for weight loss or waiting for a bariatric surgery. In the clinical sample, pregnant and lactating women were excluded.

The community sample was recruited via an online questionnaire available on social networks, via an access link on the REDCAP platform; and the clinical sample was collected at the HCPA Nutrition and Bariatric Surgery outpatient clinics and private weight loss treatment clinics. In the clinical sample, the research team contacted patients, inviting them to participate in the research, and an individual and exclusive link was generated on the platform for each invitee, which was made available only when the person agreed to participate. To minimize selection bias and to ensure that each participant answered the questionnaire only once—to avoid duplicates—a question was inserted at the end of the consent form: *“-Have you answered this questionnaire before?”* When answering yes, the questionnaire was automatically excluded. At the end of the survey, all participants who wished to, received, as a form of thanks, an online material on Mindful Eating. The study followed the conditions established in Resolution 466/12, of the Brazilian National Health Council (CNS) and was approved by the HCPA Research Ethics Committee (CAAE: 54352521.8.0000.5327 N. 2021.0447). Before entering, the survey participants were asked to confirm their eligibility and to sign an informed consent form.

To calculate the sample size, a P-Weight questionnaire, with 34 items was considered, with a 0.05 margin of error, a 95% confidence level, and a 0.85 expected Cronbach’s alpha [16]. Thus, the total sample was initially estimated at 640 individuals, 320 for the clinical sample and 320 for the community sample.

### Other Instruments

#### S-Weight

The S-Weight consists of five mutually exclusive items, among which participants had to choose in order to be allocated to one of the five stages of change (Pre-contemplative (PC) Contemplative (C) Preparation (P) Action (A) Maintenance (M), as proposed by the TTM [17].

#### Eating attitudes test-26 - (EAT-26)

This scale is used to screen for Anorexia (AN) and Bulimia (BN) (Freitas et al., 2002). In Brazil, the EAT-26 was validated in female adolescents with satisfactory psychometric properties [18]. The EAT was used in the UK and Spanish study in adults with obesity [16].

### Data Analysis

Descriptive analyses were performed using the PAWS Statistics 18 software, whereas the R software version 4.1.3 was used for exploratory (EFA) and confirmatory (CFA) factor analyses. For the EFA and CFA analyses, the total sample (n=656) and participants from the community and clinic samples were all grouped into the same group and then randomized to obtain two comparable subgroups regarding age (p=0.6) and gender (p=0.81). Following the cross-validation method used in the original articles [4,16], the Subgroup 1 (n=328) was used to perform EFA and the Subgroup 2 (n=328) for CFA. A cross-validation is desirable for EFA and CFA when the sample is large enough to randomly distribute participants into at least two groups, and it is potentially useful to perform EFA with half the sample and then use CFA to validate the factor structure with the other half [19]. The Oblimin rotation with eigenvalues >1 with extraction of four factors, and degree of item extraction >0.3 [4] was applied to the EFA for the Subgroup 1. Then, the CFA was used to the other half of the sample (Subgroup 2) to confirm the internal structure of the questionnaire. Third, internal consistency was performed for both subgroups using Cronbach’s alpha. Analysis of variance (ANOVA) was applied to the total sample (n=656) to assess the relationship between processes (dependent variables) and stages of change (independent variables) to analyze the trend in the use of processes of change. Spearman’s correlations were used to assess the convergent external validity between the P-Weight and the EAT-26.

## Results

### Participants

A total of 1,018 individuals were initially invited, of which only 656 correctly completed the questionnaire; so, the participation rate was 64.44%. Most participants (70%) were from the city of Porto Alegre. The mean age was 41.53 (SD=11.28) with BMI of 30.8 kg/m^2^ (SD=7.10). As shown in Table 1 other sample characteristics.

**Table 1.** Sociodemographic characteristics of the sample. (n=656).

| Characteristic | Community (n=330) | Clinical (n=326) | Total (n=656) |
| --- | --- | --- | --- |
| Gender, %Female | 81.5 | 74.5 | 78 |
| Age, years (a) | 41.21 (11.46) | 41.84 (11.11) | 41.53 (11.28) |
| BMI (a,b), Kg/m<sup>2</sup> | 28.41 (5.45) | 33.24 (7.71) | 30.81 (7.10) |
| %Educational level |  |  |  |
| Higher education | 63.63 | 70.85 | 67.22 |
| High School | 36.36 | 29.14 | 32.77 |
| %Skin-color |  |  |  |
| White | 85 | 83.12 | 84.29 |
| Non-white | 15 | 16.87 | 15.70 |
| %Marital status |  |  |  |
| Married | 65.45 | 59.50 | 62.5 |
| Not married | 34.54 | 40.50 | 37.5 |
| %Family income |  |  |  |
| <R\$ 7,272.00 | 66.06 | 85.27 | 75.61 |
| >R\$ 7,272.00 | 33.93 | 14.72 | 24.39 |
| a=Standard deviations |  | b=Self-reported BMI |  |

### Internal Structure

EFA was applied to Subgroup 1 (n=328). The Kaiser–Meyer–Olkin test indicated the sample adequacy for the analysis (KMO=0.92, df (528, n=328) =8,401.015; p<0.0001) and the Bartlett’s test showed a significant value with X^2^ (528) =11,809.46 and p<0.001, indicating that the correlations among the items are sufficient to perform the analysis (polycor () function of the package psych of the R programing language version 4.1.3 was used). The communalities ranged from 0.412 to 0.914, indicating a good proportion of shared variance among the analyzed items. The percentage of variance explained with four factors was 73%. The factor loadings of these items were acceptable as they reached values above 0.30, ranging from 0.374 to 1.007, as shown in Table 2.

**Table 2.**
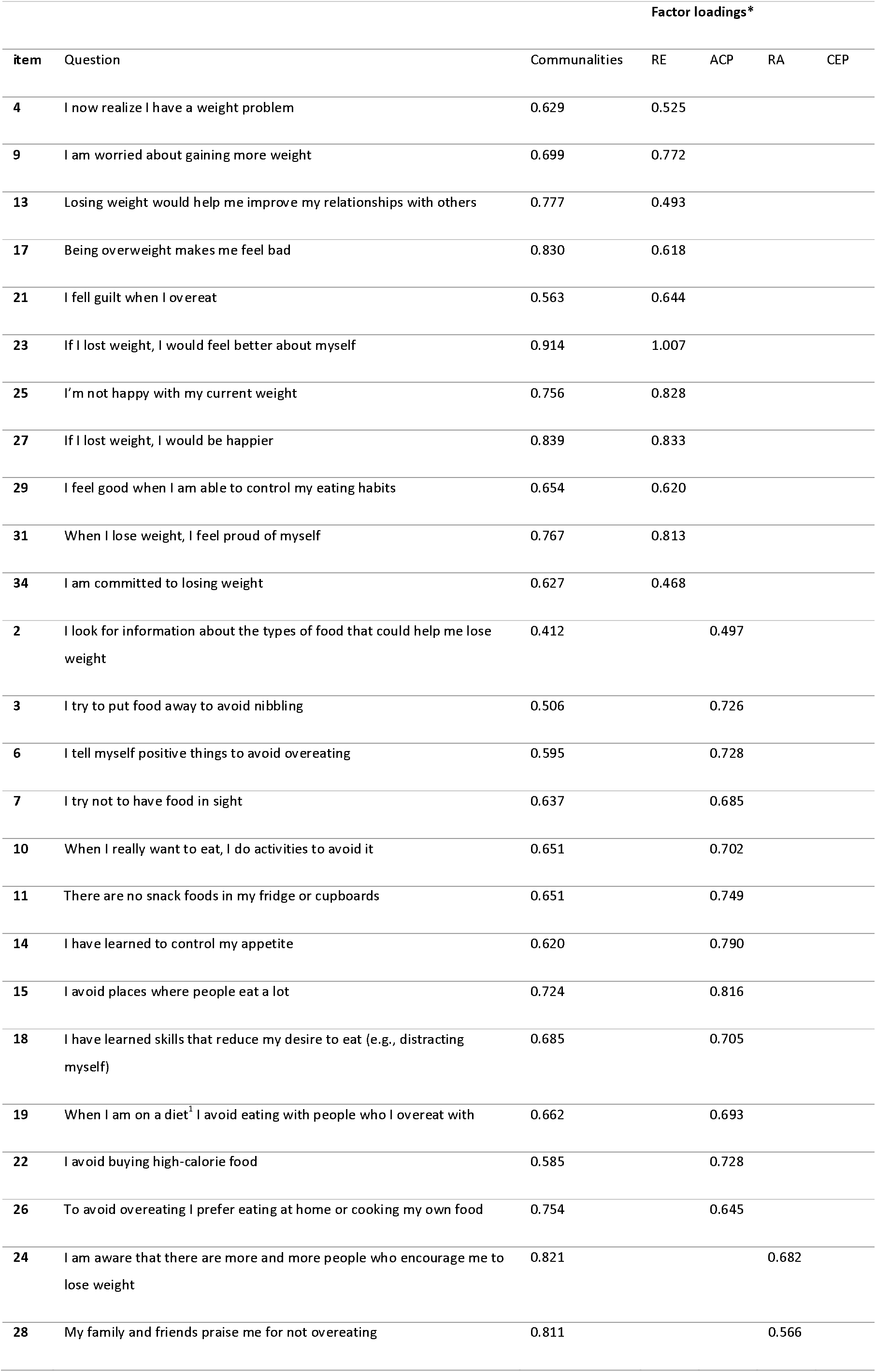
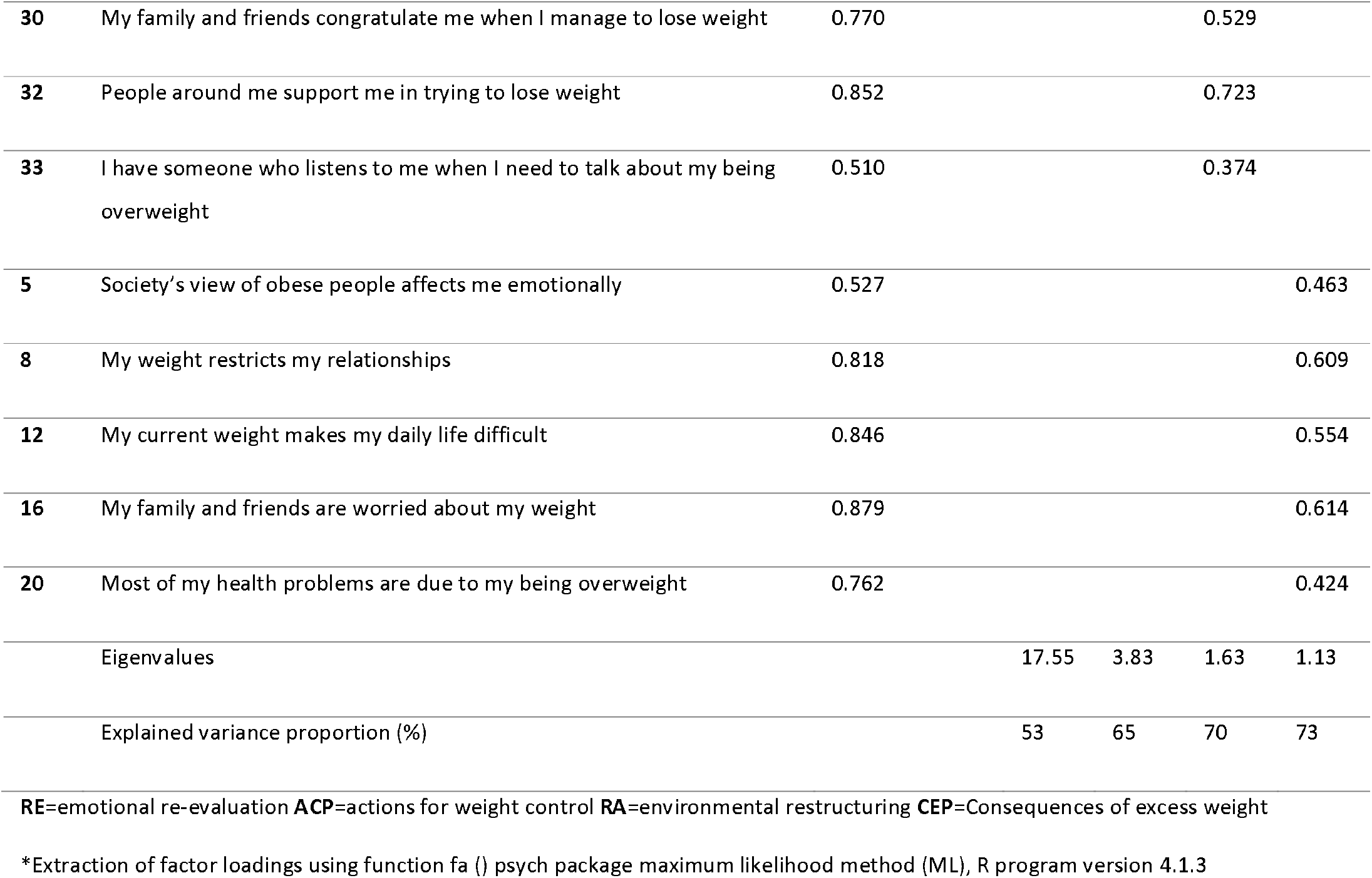
Factor loadings of P-Weight items to factors in the exploratory factor analysis (n=328).

The content analysis revealed that the items belonging to the same construct presented a slight difference in relation to the Spanish version regarding the items arrangement. The four factors revealed at EFA were labelled according to their specific content, following the same idea of the original nomenclature given by Andrés 2011: 1) Emotional Re-evaluation (RE) with 11 items; 2) Actions for weight control (ACP) with 12 items; 3) Consequences of excess weight (CEP) with five items; and 4) Environmental Restructuring (RA) with five items. One item (*“I think I should eat food with less fat”)* was excluded since the degree of extraction was less than 0.3 and the communality was less than 0.2. Therefore, the final Brazilian version of P-weight consists of 33 items. Figure 1 shows the diagram representing the four process factors of change in weight management

**Fig. 1.**
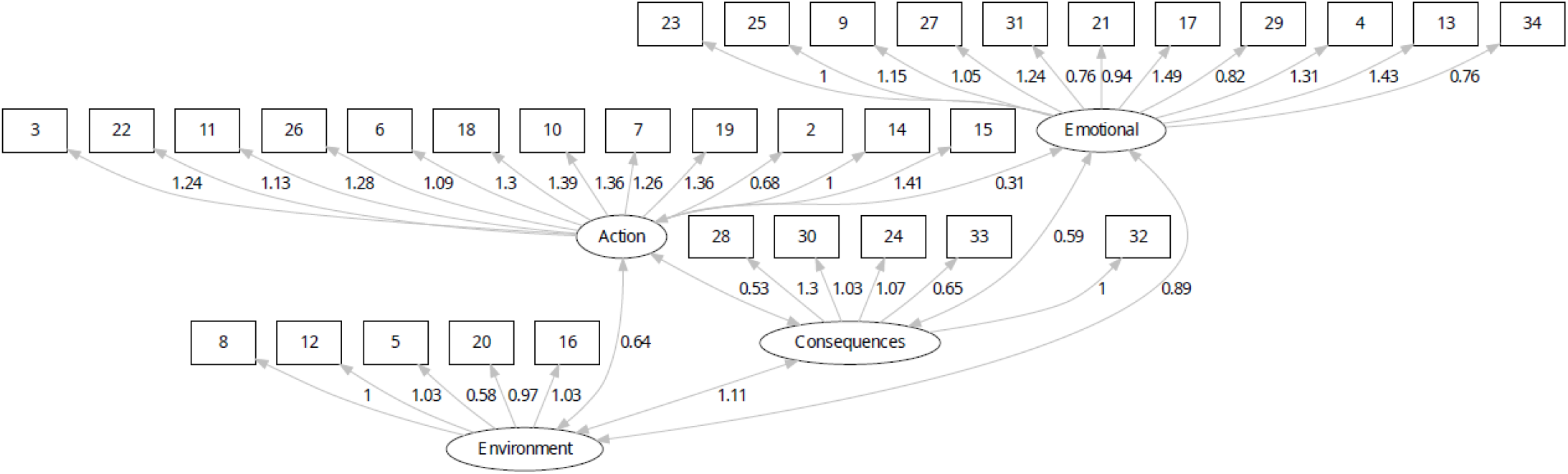
Diagram representing the four first-order factors model for processes of change in weight management. Emotional=RE=emotional re-evaluation. Action=ACP= actions for weight control. Environment=RA=environmental restructuring. Consequences=CEP=consequences of excess weight.

CFA was performed with Subgroup 2 (n=328), excluding data from Subgroup 1 for this analysis. The measurement model consisted of four factors loaded according to the results found in the EFA, showing acceptable fit of the 33 items. As shown in Table 3 the CFA structure with unstandardized parameters. The complete table is in the Supplementary Material (library function, lavaan package, R program version 4.3.1).

**Table 3.** Indices obtained in the confirmatory factor analysis.

|  | CFI | NFI/TLI | SRMR | RMSEA |
| --- | --- | --- | --- | --- |
|  | comparative fit index | normed fit index /Tucker-Lewis<br>Index | standardized root means square<br>residual | Root means square error of<br>approximation |
| Model 1 | **0.863** | **0.852** | **0.077** | **0.083** |

The Comparative Fit Index and Tucker-Lewis Index indices calculate the relative fit of the observed model when comparing it with a base model, i.e., it measures a relative improvement of the study model in relation to the standard model. The values equal to or greater than 0.90 indicate adequate adjustment, even in small samples [20]. The root mean square error of approximation is the difference index between the covariance matrix observed by degrees of freedom and the hypothetical matrix, indicating a satisfactory fit with values from 0.05 to 0.08. The standardized root mean square residual is the average of the standardized residuals between the hypothetical covariance matrices and the observed covariance; values lower than 0.10 indicates that the model is accepted [21]. As shown in Figure 1 the diagram representing the four process factors of change in weight management.

### Internal Consistency

Internal consistency was assessed for the four subdomains of P-weight derived from internal structure analysis and for the total scale in both Subgroups 1 (sample 1) and 2 (sample 2). As shown in Table 4, Cronbach’s alpha was satisfactory. Corrected item-total correlations were adequate for all items since all of them reached values over 0,30.

**Table 4.** Internal consistency analysis of the 4 factors P-Weight questionnaire in both subgroups and total sample.

|  |  | SAMPLE 1 (N=328) |  | SAMPLE 2 (N=328) |  | Total sample (N=656) |  |
| --- | --- | --- | --- | --- | --- | --- | --- |
|  | Number of Items | Cronbach's alpha | Item-total correlations (range) | Cronbach's alpha | Item-total correlations (range) | Cronbach's alpha | Item-total correlations (range) |
| RE | 11 | 0.933 | 0.558-0.827 | 0.925 | 0.541-0.803 |  |  |
| ACP | 12 | 0.922 | 0.480-0.746 | 0.921 | 0.477-0.763 |  |  |
| RA | 5 | 0.884 | 0.515-0.788 | 0.871 | 0.436-0.763 |  |  |
| CEP | 5 | 0.904 | 0.576-0.834 | 0.900 | 0.547-0.811 |  |  |
|  |  |  |  |  |  | 0.959 | 0.377 a 0.776 |

RE=emotional re-evaluation. ACP=Actions for weight control. RA=environmental restructuring. CEP=consequences of excess weight.

### Relationship between Processes and Stages of Change

As shown in Figure 2 and Table 5, ANOVA revealed significant differences in processes of change scores across the five stages of change. These differences are illustrated in the very strong linear association, with P-weight subscales scores increasing from PC to A stages of changes, except for the “consequences of excess weight (CEP)” that reached the peak in P stage. As expected, in the maintenance stage (M), the P-weight scores were lower than other stages, suggesting that the use of processes of change stabilizes or even decreases during the maintenance stage of weight management. Furthermore, in the Pre-Contemplation phase, all processes are used less than the Action stage.

**Table 5.**
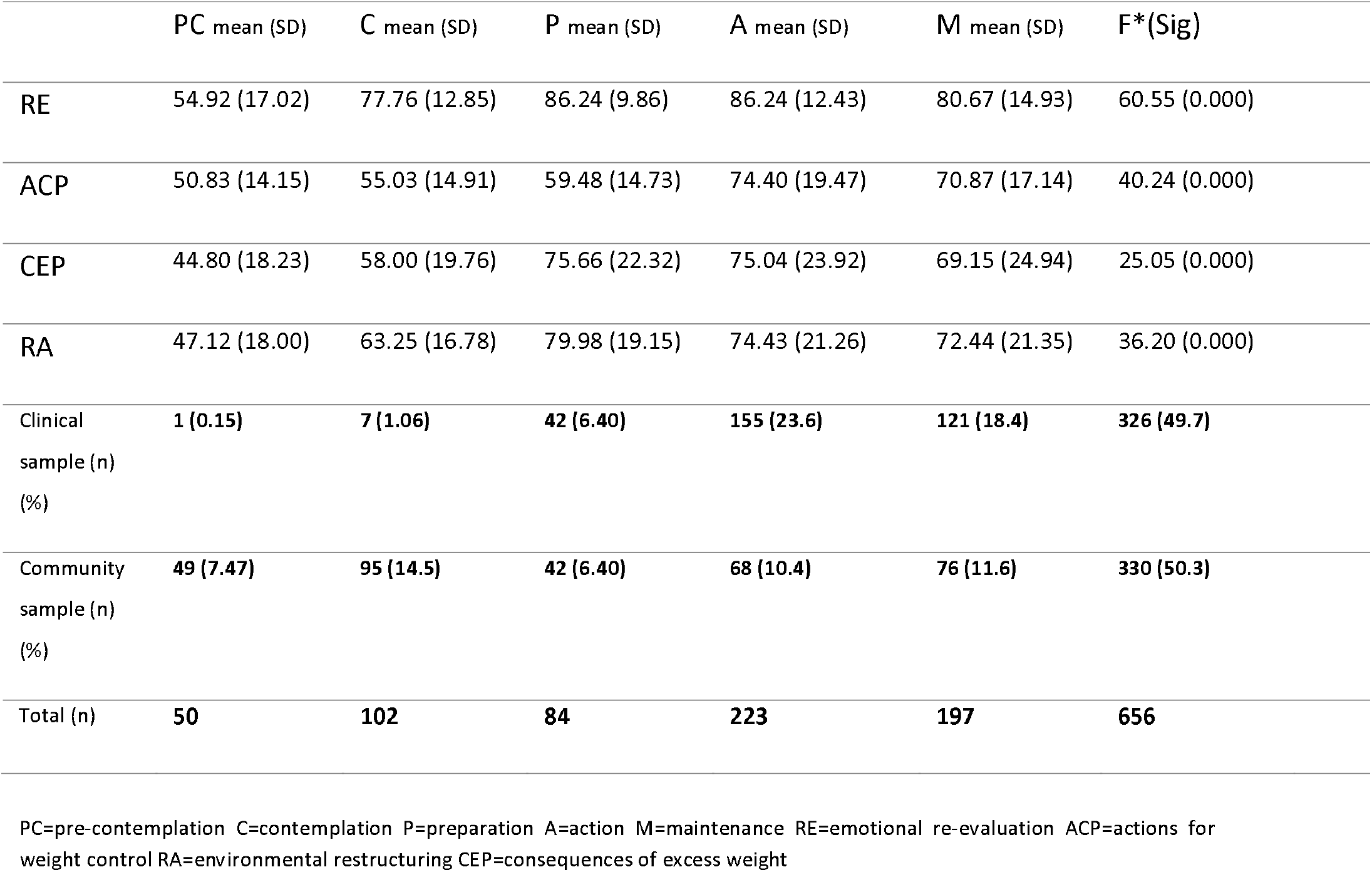
Analysis of variance of processes of change scores across stages of change (ANOVA).

**Fig. 2.**
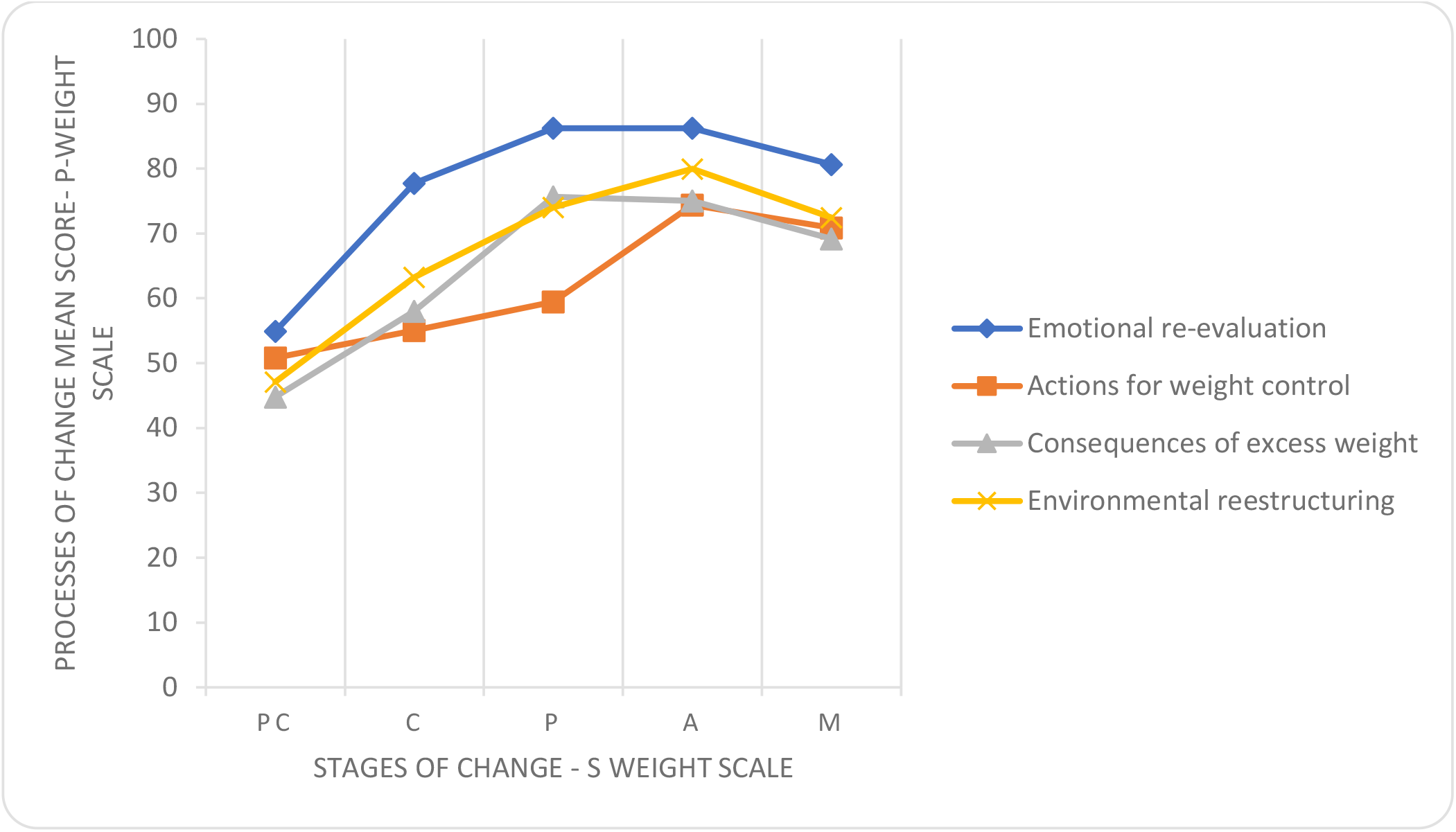
Analysis of processes of change scores across stages of change. PC=pre-contemplation; C=contemplation; P=preparation; A=action; M=maintenance.

Regarding the S-Weight, most participants (223) were in Action stage, which suggests that most of them were making efforts to manage their weight. From these, 155 participants (23.6%) were from the clinical group, whereas 68 (10.4%) participants were from the community group.

### Relationship of processes of change with other variables

As shown in Table 6, the subdomains RE, RA, ACP, and CEP of the P-Weight were significantly correlated with EAT-26, suggesting the convergent validity of the P-weight (p<0,0001).

**Table 6.** Spearman’s correlations between P-Weight and EAT-26.

|  | **RE** | **ACP** | **RE** | **CEP** |
| --- | --- | --- | --- | --- |
| **EAT 26\* – Correlation Coefficient** | 0.579 | 0.593 | 0.584 | 0.638 |
| **p (Sig)** | <0.001 | <0.001 | <0.001 | <0.001 |

RE=emotional re-evaluation. ACP=actions for weight control. RA= environmental restructuring. CEP=consequences of excess weight. \*EAT-26 =Eating attitude test-26; validated for the Brazilian population (FORTES et al., 2016)

### Sensitivity

Mean differences between clinical and community samples were calculated to assess whether the P-Weight and S-Weight were sensitive enough to detect differences between these groups. Results showed that statistically significant differences were obtained in all subscales, with participants in the clinical sample obtaining higher scores. Finally, participants from the clinical sample were allocated at more advanced stages of change for weight management. Table 7 and Figure 3 shows it.

**Table 7.** Difference means scores of the P-Weight subscales between the community and clinical sample.

| P-Weight domains | Community sample<br>(n=330) | Clinical sample<br>(n=326) | Overall sample<br>(n=656) |
| --- | --- | --- | --- |
| **RE** |  |  |  |
| Mean (SD) | 73.6 (16.4) | 88.2 (10.7) | 80.9 (15.7) |
| Median [Min, Max] | 75.4 [23.1, 100] | 89.2 [43.1, 100] | 83.1 [23.1, 100] |
| **ACP** |  |  |  |
| Mean (SD) | 56.4 (14.7) | 76.9 (17.5) | 66.6 (19.1) |
| Median [Min, Max] | 56.7 [20.0, 93.3] | 76.7 [28.3, 100] | 65.0 [20.0, 100] |
| **RA** |  |  |  |
| Mean (SD) | 58.4 (17.6) | 86.7 (14.3) | 72.4 (21.4) |
| Median [Min, Max] | 60.0 [20.0, 100] | 88.0 [36.0, 100] | 76.0 [20.0, 100] |
| **CEP** |  |  |  |
| Mean (SD) | 53.6 (19.9) | 83.4 (19.5) | 68.4 (24.7) |
| Median [Min, Max] | 52.0 [20.0, 100] | 88.0 [20.0, 100] | 72.0 [20.0, 100] |

RE=emotional re-evaluation. ACP=actions for weight control. RA=environmental restructuring CEP=consequences of excess weight

**Fig. 3.**
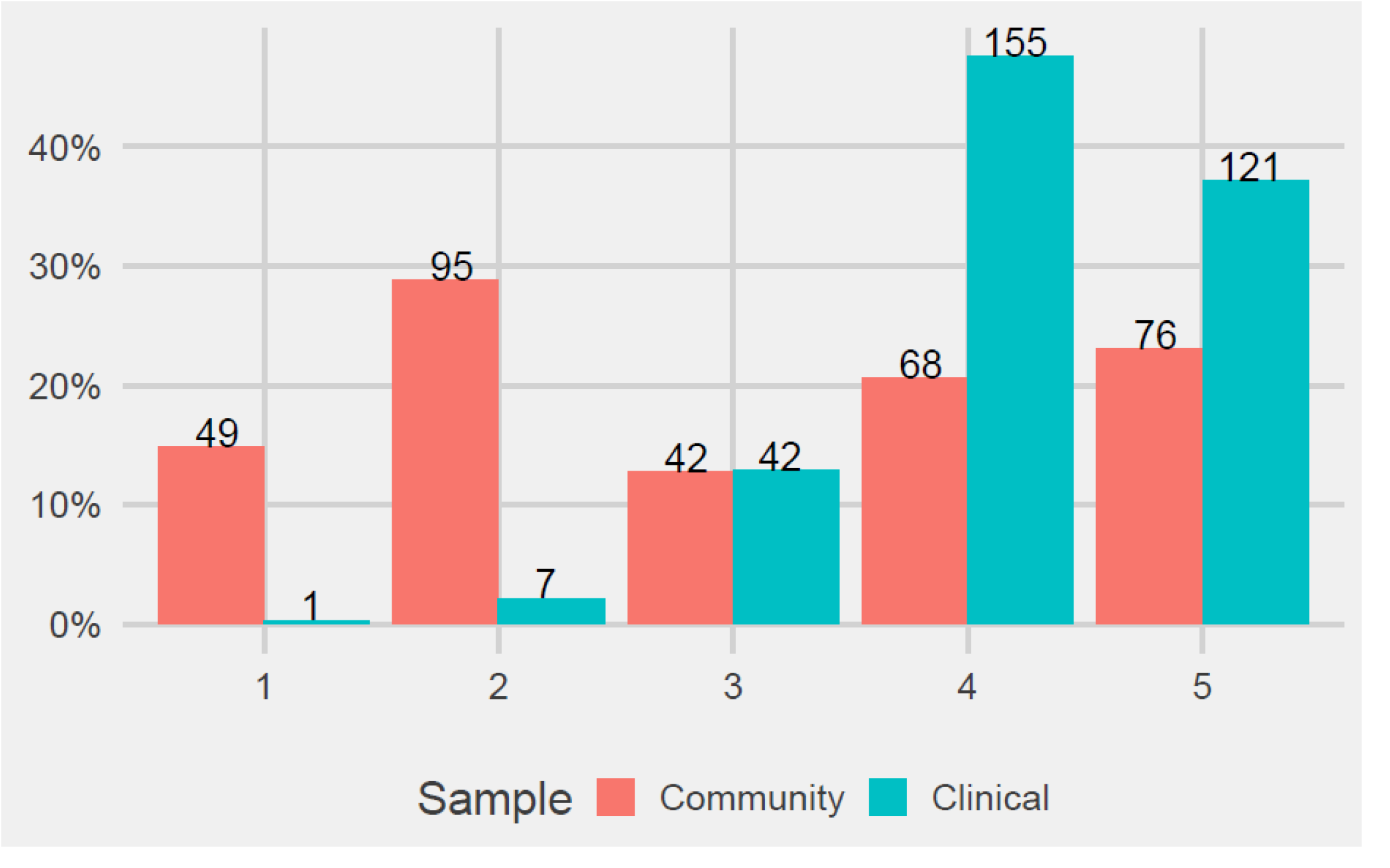
Analysis of stages of change in clinical and community samples. 1=pre-contemplation (PC) 2=contemplation (C) 3=preparation (P) 4=action (A) 5=maintenance (M). Y axis=S-Weight mean scores.

## Discussion

The Brazilian version of the P-Weight showed satisfactory psychometric properties with very high internal consistency and a positive relationship between processes and stages of change (S-Weight) and convergent validity, as indicated by a significant correlation with the EAT-26. As expected, the P-Weight mean score was higher in the clinical sample compared to the community sample, suggesting the discriminative validity of the instrument. Furthermore, the P-Weight resulted in a four-dimensional model, similar to the original Spanish version [4,16], that is, this instrument is fit to measure attitudes and behaviors involved in weight control. Therefore, the Brazilian version of the P-Weight is readily available for its use in both research and clinical practice.

Regarding internal structure, the subscales and the total scale showed adequate results, with a Cronbach’s alpha of 0.959 for the total sample, characterizing the reliability of our instrument. These results corroborate the findings in the Spanish and the UK validation studies, which showed Cronbach’s alpha of 0.960 [4] and values ranging from 0.849 to 0.955, respectively [16]. The Brazilian version of the P-Weight has 33 items, which has one item less than the original version. The item “I think I should eat food with less fat” was excluded since it has a 0.081 factor loading (less than 0.3), as recommended by Floyd and Widaman [22]. We believe that this happened due to different eating patterns between Europe and Latin America. In Brazil, people present a high consumption of fast-assimilated carbohydrates and low consumption of fatty foods, although carbohydrates- and sugar-rich foods are often also fatty foods. However, it is possible that respondents have not been aware about this technical information, which leads to a misunderstanding of this question.

Factorial analysis identified and confirmed that a model of four correlated processes of change was the most appropriate to explain behavior change in weight management. The four following subscales of the P-Weight obtained were: *Emotional re-evaluation (RE), Action for weight control (ACP), Consequences of excess weight (CEP)*, and *Environmental restructuring (RA)*. Compared with the original Spanish version, the number of items in each domain was similar with some exchange of items between domains, which was also observed in the UK validation study [16]. For example, the question “I try to put food away to avoid nibbling” in the original Spanish version is in the Environmental restructuring (RA) domain and in the UK and Brazilian versions was rearranged to the Actions for weight control (ACP) domain. This is justified because it is a specific Action item, which is taken by the individual to lose weight and, at the same time, it is related to the environment in which the individual is inserted. Likewise, the question “I now realize I have a weight problem,” which in the Spanish and the UK versions is included in the Consequences of excess weight (CEP) domain, in the Brazilian version, was rearranged to the Emotional Revaluation (RE) domain; this can be explained because besides its representation as a consequence, this item also represents the individual’s feeling when they understand that they have a weight problem. Therefore, an item can either be in the RE or RA domain or in the CEP or ACP domain since they are closely related and focus on the actions that individuals take when trying to lose weight, how they feel in this context, how the environment influences in this process, and what they understand about the effects generated by excess weight. Thus, 12 items were rearranged in different domains in comparison to previous works; all of them were assessed by the research group that considered this version satisfactory since the subscales are strongly related to each other.

The relationship between the processes and the stages of change was also a noteworthy finding. We observed a gradual increase in the use of processes of change as individuals moved from pre-contemplation (PC) to action (A) stages. People who were in PC stage used fewer processes of change compared to people in later stages. The highest use of processes of change was observed in the action (A) stage, before declining in the maintenance (M) stage. These results are in line with Andrés’s study that demonstrated that the use of processes of change was lower in the PC stage compared to subsequent stages [4]. Our results also corroborate the findings of a 12-week randomized controlled trial that concluded that the RA, CEP and ACP processes are higher in the Action stage compared to the previous stages [15]. Another recent study, using the S-Weight and P-Weight scales, revealed that patients with higher scores in the RE and ACP subscales are more likely to be in the Action stage for weight loss [23].

These findings suggest that when individuals shift their focus from *trying to lose weight* to *trying to maintain weight*, they invest less effort in processes of change. It is also possible that individuals who are closer to their ideal body weight have improved self-esteem and, therefore, remain in the Pre-Contemplation stages [13]. However, this finding may partly explain the high rates of weight regain observed in long-term studies for obesity.

Furthermore, we found a significant relationship between the four subscales of the P-Weight and measures of concern about thinness through the EAT-26, which supports the convergent validity of the scale. As expected, participants who achieved higher scores on using processes of change also achieved higher scores on external measures. These results are in line with previous findings that showed a correlation between P-Weight and EAT-40 among university students and general population [4] that participated in a program for weight loss.

The Transtheoretical Model has been used to improve changes in health-related behaviors since the 1980s, including smoking cessation and, more recently, weight management [6,24]. Thus, interventions designed to encourage behavior change are more effective when tailored to the individual’s stage of change [8]. Measuring an individual’s readiness for change in the weight management setting before undertaking an intervention can increase the effectiveness of weight loss interventions, especially in the long term [25]. Therefore, it is crucial to have reliable and validated measurement tools to assess the processes and stages of behavior change in weight management. The Brazilian version of the P-Weight has shown adequate psychometric properties and can be a useful tool to identify the individual’s behavior change processes in relation to their weight. Personalized interventions that promote specific change processes and encourage progress through the motivational stages can aid in weight management.

This study has some limitations. Firstly, the non-random sampling procedure may not accurately represent the Brazilian population, so the results should not be generalized to populations with different characteristics. Secondly, participants’ self-reported weight and height may be biased and could affect questionnaire responses. Finally, we highlight that the correlation between P-Weight scores and EAT-26 should be viewed as evidence of the scale convergent validity, and not as a measure of mental health issues.

## Conclusion

The results of this study show that the Brazilian version of the P-Weight has satisfactory psychometric properties, making it a valuable application for weight loss interventions based on the Transtheoretical Model (TTM). The use of motivational strategies is crucial to promote adherence to treatments for overweight and obesity. The availability of validated instruments that allow the evaluation of change processes, based on the TTM, is a great advance and should contribute enormously to this important public health problem. We highlight that individualizing weight management interventions according to each patient’s motivational stage and processes of change allows for health professionals to employ more effective treatments according to each individual’s motivational reality.

## Data Availability

All data produced in the present study are available upon reasonable request to the authors

## Statements

## Acknowledgments

The authors thank to the Higher Education Personnel Improvement Coordination - Brazil (CAPES), National Council for Scientific and Technological Development and Research Incentive Fund of the Clinical Hospital of Porto Alegre for their funding support (FIPE/HCPA 2021.0447). Adriane R. Rosa would like to thank CNPq, PQ number 305707-2015/09.

## Statement of Ethics

The study followed the conditions established in Resolution 466/12, of the Brazilian National Health Council (CNS) in accordance with the World Medical Association Declaration of Helsinki.

### Study approval statement

This study protocol was reviewed and approved by the HCPA Research Ethics Committee, approval number (CAAE: 54352521.8.0000.5327 N. 2021.0447).

### Consent to participate statement

Before entering, the survey participants were asked to confirm their eligibility and to sign an informed consent form. The study followed the bioethical principles of autonomy, beneficence, non-maleficence, veracity and confidentiality. All participants were informed about the risks related to psychological discomfort when answering questions about eating habits, their relationship with food and their body weight. To minimize these discomforts, subjects were informed that they could discontinue their participation in the study at any time, since participation is voluntary. To minimize the chance of breach of confidentiality, the questionnaires were anonymous. In addition, we use a specific tool for this type of study, which is the REDCAP. The completed online questionnaires were kept electronically and confidentially for a period of 5 years. All participants completed the free and informed consent form (TCLE) online and only after that the research started and they were free to discontinue the study at any time without this implying any harm to the participant. The research group declares to know and comply with the requirements of the General Data Protection Law (Law No. 13,709, of August 14, 2018) regarding the processing of personal data and sensitive personal data that will be used for the execution of this research project.

## Author Contributions

ARR and QC conceived the manuscript idea. QC and BBM performed the statistical analysis. PRZ, BBM, JFG, and QC wrote the manuscript. ARR supervised the findings of this work. All authors discussed the results and contributed to the final manuscript.

## Data Availability Statement

All data generated or analyzed during this study are included in this article. Further enquiries can be directed to the corresponding author.

